# No Correlation of Structural Anterior-Segment OCT Biomarkers with Bleb Vessel Density and Surgical Success after Preserflo Microshunt Implantation

**DOI:** 10.1101/2024.06.30.24309720

**Authors:** Martin Kallab, Sarah Hinterberger, Sophie Schneider, Olivia Murauer, Anna-Sophie Reisinger, Susanne Strohmaier, Alex S. Huang, Matthias Bolz, Clemens A. Strohmaier

## Abstract

**Purpose:** To evaluate anterior segment optical coherence tomography (AS-OCT) parameters of bleb wall thickness (BWT) and total bleb height (TBH) in the early postoperative phase after Preserflo Microshunt (PM) implantation for their correlation to (a) surgical revision and (b) AS-OCT angiography (AS-OCTA) derived bleb vessel density (BVD).

**Methods:** A total of 23 patients with pharmacologically uncontrolled open angle glaucoma were studied. Post-operatively (at 1, 2, and 4 weeks and 2, 3, 6, 9, and 12 months) AS-OCT measurements of BWT/TBH and AS-OCTA measurements of BVD were acquired. Surgical revisions (needling or open revision) were recorded. Correlations of BWT and TBH to (a) need for surgical revision and to (b) BVD were assessed.

**Results:** In 10 of 23 patients, surgical revisions were performed 4 to 48 weeks after PM implantation. At 1, 2, and 4 weeks after surgery neither BWT nor TBH were significantly associated with future surgical revisions (BWT/TBH p-values: 1W 0.217/0.878, 2W 0.670/0.528, 4W 0.171/0.430). No correlations between BWT or TBH and BVD were found for any evaluated timepoint (1W, 2W, 4W).

**Conclusions:** Structural AS-OCT parameters were not predictive of the need for surgical revision after PM implantation. Consistent with this finding, these parameters were also not correlated with AS-OCTA derived BVD, which was shown to be a good biomarker for failure in a previous analysis. The discrepancy to similar studies after trabeculectomy may be due to bleb drainage differences between TE and PM. BVD seems to be a better predictor of surgical revision after PM implantation than BWT/TBH.

## Introduction

Lowering intraocular pressure (IOP) is still the mainstay of glaucoma treatment. Surgical options to lower IOP have seen numerous advancements towards a more standardized and probably safer approach to filtration surgery. Still, a considerable number of patients need secondary interventions (1) and therefore an intensified follow-up regime is required for optimal patient care (2).

Filtration surgeries aim at generating an aqueous humor bypass from the anterior chamber to the subconjunctival/subtenon space either by creating a surgical scleral flap (trabeculectomy (TE)) or through implant-assisted procedures such as Xen gel (XG) (3, 4) or more recently Preserflo Microshunt (PM) (5, 6). While TE consistently shows greater IOP lowering efficacy than PM implantation, PM is still associated with marked IOP reduction that is close to TE and demonstrates a favorable peri- and postoperative risk profile.(7–9) Therefore, PM is increasingly deployed as a first-line glaucoma surgery as opposed to traditional TE.(7–9)

Filtration surgeries in general depend on proper filtration bleb (FB) function and, therefore, FB evaluation, either by clinical observation or by anterior segment imaging, plays a central role in postoperative patient management. The value of clinical bleb grading scales, e.g. the Moorfields Bleb Grading System (MBGS)(10) and Indiana Bleb Appearance Grading Scale (IBAGS)(11), is, however, impaired by partly subjective grading algorithms(12, 13). They also lack detailed evaluation of internal bleb structure and dimensions. To overcome these limitations, anterior segment optical-coherence tomography (AS-OCT) has been utilized to develop objective biomarkers for bleb function.(14) Nowadays, bleb wall thickness (BWT) and total bleb height (TBH) are among the most frequently studied quantitative AS-OCT-based structural bleb parameters and have been repeatedly shown to correlate well with bleb function after TE.(15–22) Results looking at the same endpoints after PM implantation have been equivocal.(23–25)

Moreover, while existing cross-sectional data on correlation between concomitantly recorded AS-OCT parameters and IOP undoubtedly help to extend our knowledge on pathophysiological processes linked to bleb failure comprising vascularization, fibrosis and bleb encapsulation, their utility in clinical routine care may, however, be limited. From a clinical perspective, early biomarkers informing the clinician about future bleb failure risk are needed to allow a personalized approach to glaucoma patient care. For the above-mentioned quantitative AS-OCT based bleb parameters, information on their predictive value is, however, scarce after TE (15, 16) and not available after PM implantation. As we could recently identify AS-OCT angiography (AS-OCTA) measured bleb vessel density (BVD) two and four weeks after surgery as a predictor for surgical revision up to one year after PM primary surgery (26), we set out to investigate this relationship for structural AS-OCT bleb parameters as well.

Therefore, the aim of the current analysis was to evaluate structural AS-OCT bleb parameters, BWT and TBH, in the early postoperative phase concerning their predictive value for surgical revision after PM implantation in a longitudinal study cohort and to correlate those parameters with AS-OCTA derived bleb vascularity information.

## Methods

### Study design and patient selection

The protocol of this single-center study (Department of Ophthalmology and Optometry, Kepler University Hospital, Linz, Austria) was approved by the Ethics Committee of the Johannes Kepler University (EC-No.: B-142-17). Written informed consent was obtained from every patient before study inclusion and the tenets of the Declaration of Helsinki were followed during all study-related procedures.

Inclusion criteria included glaucoma type (primary open angle glaucoma, pseudoexfoliation glaucoma, or pigment dispersion glaucoma) and indication for PM implantation (maximal tolerated IOP-reducing medical therapy AND uncontrolled IOP > 21 mmHg and/or visual field (VF) progression as tested using the 30-2 SITA fast algorithm of Humphrey Field Analyzer II 750 ((Carl Zeiss Meditec Inc., Dublin, CA, USA) and/or progressive retinal nerve fiber layer thickness reduction as measured using Spectralis OCT (Heidelberg Engineering, Heidelberg, Germany)). Exclusion criteria included, angle closure glaucoma and previous filtration surgery.

### Preserflo Microshunt: medical device and implantation surgery

Descriptions of in-depth surgical technique specifications of the PM medical device (Santen Pharmaceutical, Osaka, Japan) have been published elsewhere.(5, 6) Briefly, the PM glaucoma drainage device is made from poly(styrene-block-isobutylene-block-styrene) (SIBS), a highly biocompatible and bioinert material, and has following basic dimensions: length: 8.5mm, outer diameter: 350 µm, inner diameter: 70 µm. In the European Union the PM is CE-marked and approved for use as glaucoma drainage device, FDA approval is pending.

In this study, standard Mitomycin C (MMC) augmented PM implantation was performed in the supero-temporal or supero-nasal quadrant under subtenon or general anesthesia. MMC-soaked sponges (0.2 mg/ml) were applied for 3 minutes after dissection of conjunctiva before the scleral pocket and tunnel to the anterior chamber were created. Then, the PM was placed in the tunnel with its fins inside the scleral pocket. Finally, flow through the implant was checked by balanced saline solution injection, and conjunctival sutures were placed to close all wounds.

### Postoperative treatment and examinations

Preservative-free antibiotic (Ofloxacin, 1 week) and steroid (Dexamethasone, tapered over 12 weeks) drops were prescribed according to a standardized protocol. One, two and four week(s), as well as two, three, six, nine and twelve months after surgery slit-lamp examinations, IOP measurements, structural AS-OCT measurements (Casia 2 Cornea/Anterior Segment OCT, Tomey Corporation, Nagoya, Japan) and AS-OCTA measurements (PLEX Elite 9000; Carl Zeiss Meditec, Dublin, CA, USA) were performed. Moreover, necessity and number of IOP lowering medications (MEDs) as well as surgical revisions (needling or open revision) were recorded. Experienced clinicians (S.SCH, A-S.R., CA.S.) offered a surgical revision if postoperative IOP exceeded the preoperatively defined target pressure and no bleb or a scared bleb was clinically visible. Depending on fulfilment of the endpoint “surgical revision”, which included either needling or open revision, patients were allocated to the outcome groups “surgical revision” or “no surgical revision”.

### AS-OCT scan acquisition and image analysis

All AS-OCT images were acquired using the Casia2 AS-OCT with on-board software. Operators instructed the patients to look down to expose the FB in the superior-nasal or -temporal quadrant. Upon visualization of the FB and PM in the real-time preview OCT-image, an AS-OCT volume was acquired using the standard settings of the “Bleb” imaging mode (volume size 12x12mm, number of B-scans: 256, number of A-scans per B-scan: 400). To measure BWT and TBH, a B-scan was selected based on visibility of following structures to ensure intra- and inter-individual comparability: posterior PM tip, complete bleb wall above PM, and episcleral fluid. If more than one scan fulfilled all criteria, the B-scan with the higher bleb was selected. If no episcleral fluid was visible in any scan (i.e. not present in individual bleb), this criterion was omitted.

All AS-OCTA images were acquired using the PLEX Elite 9000 OCTA in combination with a 10dpt anterior segment add-on lens. Details of our AS-OCTA image processing and BVD calculation approach were published previously.(26) In short, AS-OCTA slabs with the PM scleral passage position marked in an overlay were exported. Using Fijii ImageJ (27) motion artifacts were removed, before a circular region of interest (400px diameter) centered around the scleral passage of the PM was defined. Images were then binarized, and the BVD was calculated as the proportion of white pixels.

### Statistical analysis

If not differently indicated, all values were presented as means±standard deviation (SD). Logistic regression modelling in combination with the likelihood-ratio test was used to evaluate predictive value of independent variables (BWT or TBH) for the dependent dichotomic variable (surgical revision and no surgical revision). Pearson bivariate correlation was used to evaluate associations of structural AS-OCT parameters (BWT and TBH) with AS-OCTA parameters (BVD). Prism 10 (GraphPad Software, Boston, MA, USA) was used for statistical testing and graph production.

## Results

### Baseline characteristics

Twenty-three patients with OAG and preoperative IOP of 23.57±7.75 mmHg received PM implantation and were included in this study. In ten patients, needling or open revision were performed during the follow up period of 1 year. Earliest surgical revisions were performed after the 4 weeks’ postoperative follow-up. Further baseline characteristics in the whole cohort and the two outcome groups are summarized in Table 1.

**Table 1.** Baseline characteristics. SR: surgical revision, OAG: open angle glaucoma, POAG: primary open angle glaucoma, PDG: pigment dispersion glaucoma, PXG: pseudoexfoliation glaucoma, MD: visual field mean deviation, dB: decibel, 1W: one week

| Table 1. Baseline characteristics. |  |  |  |
| --- | --- | --- | --- |
|  | Whole cohort | SR | no SR |
| N | 23 | 10 | 13 |
| Age (mean±SD) | 69.26±10.05 | 68.2±3.99 | 70.08±13.09 |
| Sex (f/m) | 13/10 | 5/5 | 8/5 |
| OAG type<br>(POAG/PDG/PXG) | 17/1/5 | 9/0/1 | 8/1/4 |
| MD (dB, mean±SD) | -9.83±7.43 | -6.13±5.74 | -12.67±7.52 |
| IOP-preOP | 23.57±7.75 | 24.40±8.80 | 22.92±7.15 |
| MEDs-preOP<br>(mean±SD) | 2.96±1.11 | 3.40±0.52 | 2.62±1.33 |
| IOP-postOP (1W) | 8.30±2.12 | 9.60±1.84 | 7.31±1.80 |
| Timepoint of SR<br>(week, median, min-max) | - | 10, 4-48 | - |

### Descriptive statistics of postoperative IOP, BWT and TBH

Postoperative IOP one, two and four week(s), as well as two, three, six, nine and twelve months after surgery were 8.30±2.12, 9.17 ±2.33, 11.70±4.39, 13.48±5.83, 11.87 ±4.49, 12.30±6.65, 11.87±3.11 and 13.05±4,12 mmHg. Postoperative BWTs and TBHs over time are summarized in Table 2 and Figure.

**Figure.**
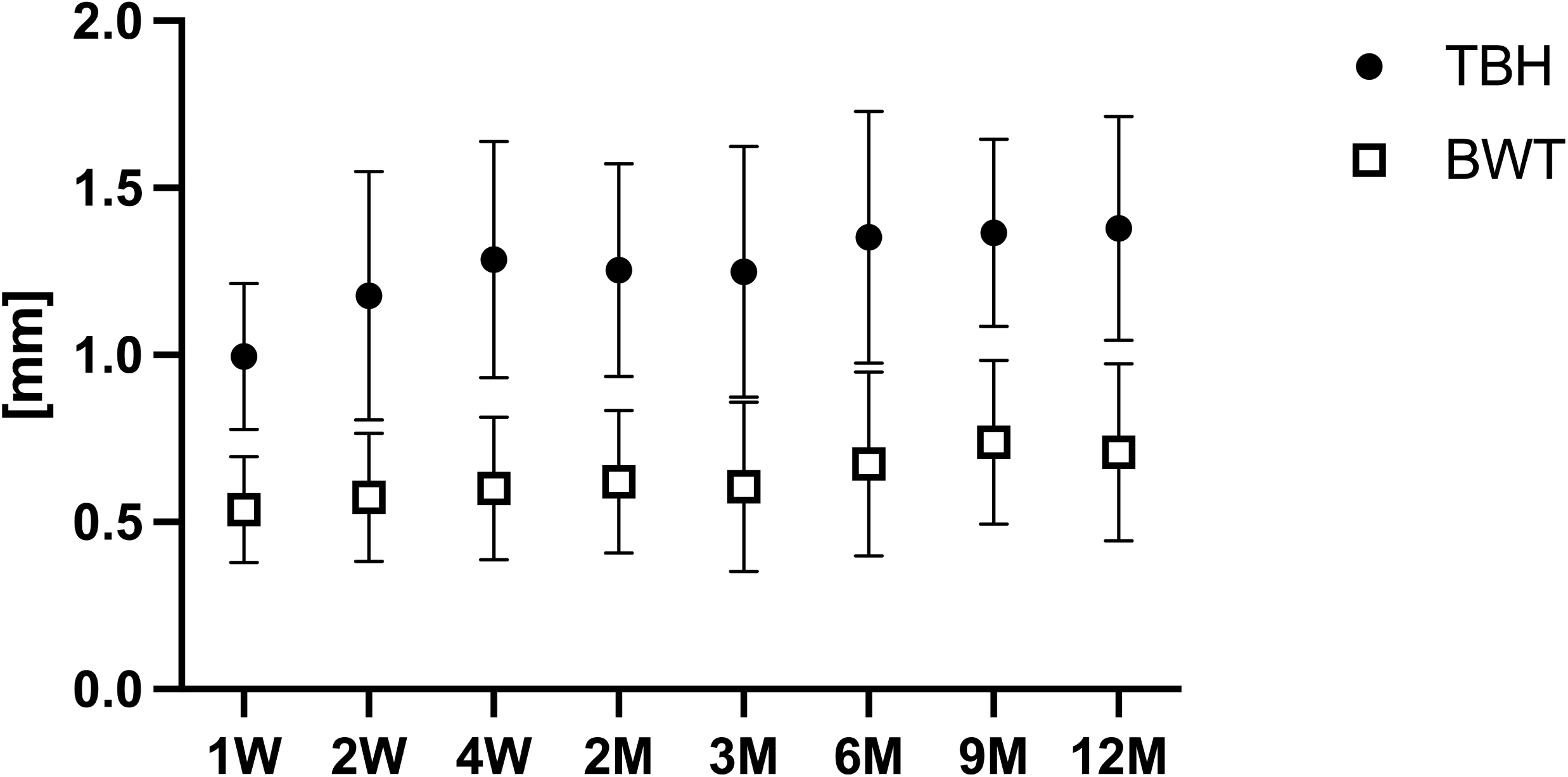
Time Course of bleb wall thickness (BWT) and total bleb height (TBH). TBH: total bleb height, BWT: bleb wall thickness.

**Table 2.** Tabulated time course of TBH and BWT values. TBH: total bleb height, BWT: bleb wall thickness

| Postoperative<br>Timepoint | TBH [$\mu\text{m}$] | BWT [$\mu\text{m}$] |
| --- | --- | --- |
| 1 week | 996±218 | 538±159 |
| 2 weeks | 1177±373 | 574±192 |
| 4 weeks | 1286±354 | 601±214 |
| 2 months | 1255±319 | 621±213 |
| 3 months | 1249±375 | 607±254 |
| 6 months | 1353±377 | 674±276 |
| 9 months | 1366±281 | 739±246 |
| 12 months | 1379±335 | 709±266 |

### Association of BWT and TBH with future surgical revision

As earliest surgical revisions were performed after the 4 weeks’ postoperative follow-up, 1,2 and 4 weeks’ BWT and TBH were evaluated. Upon logistic regression modelling, both, BWT and TBH, 1, 2 and 4 weeks after surgery were not found to be significantly associated with future surgical revisions (BWT/TBH likelihood-ratio test p-values: 1W 0.2169/0.8776, 2W 0.6704/0.5281, 4W 0.1706/0.4298). Areas under the receiver operating characteristics curves (AUROCs) were low for both parameters at all evaluated timepoints (BWT/TBH AUROCSs 1W: 0.612/0.519, 2W: 0.539/0.581, 4W: 0.631/0.585).

### Correlation between structural (BWT, TBH) and vascular (BVD) data

As we could recently show, that AS-OCTA measured BVDs 2 and 4 weeks after PM implantation are predictive for future surgical revision (26), correlations between structural parameters (BWT and TBH) and bleb vascularity (BVD) were evaluated. No significant correlations could be found for any timepoint or parameter (Pearson r between 0.339 and -0.170, p >0.05 for all correlations). Detailed results are summarized in Table 3.

**Table 3.** Correlations between AS-OCT derived structural and AS-OCTA derived vascular bleb parameters. BWT: bleb wall thickness, BVD: bleb vessel density, TBH: total bleb height

| Variables | Pearson r | P Value |
| --- | --- | --- |
| 1W BWT / 1W BVD | 0.339 | 0.1553 |
| 1W TBH / 1W BVD | 0.191 | 0.4330 |
| 2W BWT / 2W BVD | 0.119 | 0.6383 |
| 2W TBH / 2W BVD | -0.170 | 0.4990 |
| 4W BWT / 4W BVD | -0.081 | 0.7409 |
| 4W TBH / 4W BVD | -0.077 | 0.7527 |

## Discussion

In this study we analyzed the predictive value of two quantitative, AS-OCT derived bleb parameters (BWT and TBH) in the early postoperative phase for surgical revision after PM implantation. Both, BWT and TBH, did not predict surgical revisions after PM implantation and did not correlate with AS-OCTA-measured bleb vascularity (BVD) at 1, 2 or 4 weeks after primary surgery. The latter analysis was performed because we found BVD to be an early predictive marker for future surgical revision up to one year after PM implantation in the same cohort.(26)

In an effort to find objective markers for bleb function AS-OCT has become an increasingly popular tool both in clinical research and routine care.(14) Concerning bleb evaluation after PM implantation, 3 studies have evaluated correlations of quantitative bleb markers with IOP and obtained inconsistent results.(23–25) In 2022 Ibarz Barbera et. al found no correlations between IOP and horizontal or vertical (TBH minus BWT) bleb dimensions in a longitudinal study with up to 3 months follow up.(25) The same group later analyzed various bleb parameters in a cross-sectional design 1 year after PM implantation and reported a significant correlation of TBH and BWT with surgical success (defined by IOP between 6 and 17 mmHg and >=20% reduction without medication) in univariate analysis, which did not persist upon multivariate analysis.(24) Finally, in a retrospective cross-sectional analysis, Gambini et al. did not find associations between bleb dimensions and IOP 6 months after PM implantation.(23)

The course of BWT and TBH in our cohort is, however, comparable to published values by Ibarz Barbera et al. after PM implantation with a noticeable increase of both parameters in the first 3-6 months, transitioning to a plateau-like phase in the second half of the one year follow-up period.(24)

The available body of evidence for predictive AS-OCT biomarkers after TE is in contrast to our results for PM. Narita et al. found TBH and BWT after 2 weeks to be predictive for surgical success (defined by an IOP <= 15 mmHg and >20% reduction without surgical revision) up to one year after surgery.(16) Waibel et al. detected higher BWT as early as one week and higher bleb cavity height (TBH minus BWT) two weeks after TE in functioning blebs as compared to non-functioning blebs in a follow-up period of three months.(15) These data are backed by further studies showing correlations (at the same timepoints) between quantitative AS-OCT bleb parameters and IOP (or IOP-based success endpoints) after TE.(17–22)

A possible reason for the discrepancy between TE and PM, could be differences in bleb position and morphology as recently detected by Hasan et al. in an extensive comparative study of functioning blebs after TE and PM implantation.(28) Compared to TE blebs, PM blebs showed reduced conjunctival microcysts, a more consistent Tenon appearance and a more posterior drainage (through posterior PM tip where Tenon’s capsule is thicker). Based on these findings the authors suggested bleb drainage differences between the two surgical techniques.(28)

In this context it appears also notable that the vascular parameter BVD, which is independent of bleb thickness, was recently found to be an early predictive marker for surgical revision after PM implantation by our research group.(26) This result agrees with AS-OCTA-based vascularity biomarkers after TE (29–31) which could be indicative for a more procedure-independent validity of bleb vascularity biomarkers.

Studies correlating or combining, structural AS-OCT-based and vascular AS-OCTA-based bleb parameters are rare and currently available for TE and XG implantation but not for PM implantation. Yin et al. found a correlation between vessel area and TBH 1 month after TE.(31) Hayek et al. analyzed preoperative conjunctival vessel density and found an association with postoperative microcysts.(30) Six months after XG implantation, Mastropasqua et al. detected a correlation of TBH and bleb wall microcysts with vessel displacement area, a marker for flow voids.(32) In an effort to combine structural and angiographic data Luo et al. designed an objective bleb evaluation score in which information on vessel density, bleb height and microcysts were included.(33)

To our knowledge the current study is the first to correlate AS-OCT-based structural and AS-OCTA-based vascular bleb parameters after PM implantation. While these correlations are all statistically insignificant, this result does not come as a surprise as BVD at two of the three presented timepoints (postoperative week two and four) were recently shown to be an early predictor of surgical revision after PM implantation (26) and BWT/TBH, as presented in this study, are not. Discrepancy of BWT/TBH data between TE and PM implantation, as above extensively discussed, might also explain diverging results between the study of Yin et al. and our analysis.(31)

The present study has several strengths and limitations. Concomitant presentation and correlation of AS-OCT- and AS-OCTA-based parameters, the prospective study design and the concise, clinically relevant endpoint definition are strengths of this study. While the two earlier points have already been thoroughly discussed, the latter shall also be briefly outlined. We deliberately refrained from including IOP to our endpoint definition, which is commonly used in comparable studies. Instead, we relied solely on the clinically relevant necessity for surgical revision, which is related to both IOP and clinical bleb appearance. This was done to avoid misclassification as bleb failure solely based on concepts such as threshold IOP or percent IOP reduction as both may be arbitrary and definitely depend on baseline IOP.

An obvious limitation of this study is its rather small sample size, especially as we present not significant, negative results and draw conclusions. The sample size in the present study would, however, at least have allowed for the detection of an association if the min/max values for either parameters modified this risk by 20%, assuming a baseline surgical revision incidence of 30%. Furthermore, we consider the type II error risk to be low due to following two circumstances: 1) AS-OCT studies after TE with comparable sample sizes detected differences in bleb thickness parameters.(15, 19) 2) We recently found BVD two and four weeks after PM implantation to be a predictor for surgical revision in a study with comparable sample size.(26)

In conclusion, structural AS-OCT-based bleb thickness parameters (BWT and TBH) in the early postoperative phase, were not found to be predictive for surgical revision after PM implantation and did not correlate with AS-OCTA-derived BVD. Discrepancy to the value of quantitative AS-OCT parameters after TE and recently published AS-OCTA-derived vascular parameters after PM implantation may be associated with differences in bleb drainage between TE and PM. AS-OCTA-derived BVD seems to be a better predictor of surgical revision after PM implantation than BWT/TBH.

## Data Availability

All data produced in the present study are available upon reasonable request to the authors.

## Acknowledgements

This study was funded by NIH grant R01EY030501 (ASH) and an unrestricted grant from Santen SA. Santen SA was not involved in study design and conduct, data analysis and interpretation or manuscript production.

## Financial disclosure

CAS: Santen(Consultant, Research Support), Zeiss(Honorarium), AbbVie(Consultant, Honorarium), Elios Vision (Consultant, Honorarium).

ASH: Allergan(C), Amydis(C), Celanese(C), Diagnosys(F), Equinox(C), Glaukos(C,F), Heidelberg Engineering(F), QLARIS(C), Santen(C), Topcon(C).

## References

1. Marolo P, Reibaldi M, Fallico M, Maugeri A, Barchitta M, Agodi A, et al. Reintervention rate in glaucoma filtering surgery: A systematic review and meta-analysis. Eur J Ophthalmol. 2022;32(5):2515–31.

2. Marquardt D, Lieb WE, Grehn F. Intensified postoperative care versus conventional follow-up: a retrospective long-term analysis of 177 trabeculectomies. Graefes Arch Clin Exp Ophthalmol. 2004;242(2):106–13.

3. Schlenker MB, Gulamhusein H, Conrad-Hengerer I, Somers A, Lenzhofer M, Stalmans I, et al. Efficacy, Safety, and Risk Factors for Failure of Standalone Ab Interno Gelatin Microstent Implantation versus Standalone Trabeculectomy. Ophthalmology. 2017;124(11):1579–88.

4. Lewis RA. Ab interno approach to the subconjunctival space using a collagen glaucoma stent. J Cataract Refract Surg. 2014;40(8):1301–6.

5. Pinchuk L, Riss I, Batlle JF, Kato YP, Martin JB, Arrieta E, et al. The use of poly(styrene-block-isobutylene-block-styrene) as a microshunt to treat glaucoma. Regen Biomater. 2016;3(2):137–42.

6. Pinchuk L, Riss I, Batlle JF, Kato YP, Martin JB, Arrieta E, et al. The development of a micro-shunt made from poly(styrene-block-isobutylene-block-styrene) to treat glaucoma. J Biomed Mater Res B Appl Biomater. 2017;105(1):211–21.

7. Batlle JF, Fantes F, Riss I, Pinchuk L, Alburquerque R, Kato YP, et al. Three-Year Follow-up of a Novel Aqueous Humor MicroShunt. J Glaucoma. 2016;25(2):e58–65.

8. Baker ND, Barnebey HS, Moster MR, Stiles MC, Vold SD, Khatana AK, et al. Ab-Externo MicroShunt versus Trabeculectomy in Primary Open-Angle Glaucoma: One-Year Results from a 2-Year Randomized, Multicenter Study. Ophthalmology. 2021;128(12):1710–21.

9. Beckers HJM, Aptel F, Webers CAB, Bluwol E, Martinez-de-la-Casa JM, Garcia-Feijoo J, et al. Safety and Effectiveness of the PRESERFLO(R) MicroShunt in Primary Open-Angle Glaucoma: Results from a 2-Year Multicenter Study. Ophthalmol Glaucoma. 2022;5(2):195–209.

10. Wells AP, Crowston JG, Marks J, Kirwan JF, Smith G, Clarke JC, et al. A pilot study of a system for grading of drainage blebs after glaucoma surgery. J Glaucoma. 2004;13(6):454–60.

11. Cantor LB, Mantravadi A, WuDunn D, Swamynathan K, Cortes A. Morphologic classification of filtering blebs after glaucoma filtration surgery: the Indiana Bleb Appearance Grading Scale. J Glaucoma. 2003;12(3):266–71.

12. Hoffmann EM, Herzog D, Wasielica-Poslednik J, Butsch C, Schuster AK. Bleb grading by photographs versus bleb grading by slit-lamp examination. Acta Ophthalmol. 2020;98(5):e607–e10.

13. Wells AP, Ashraff NN, Hall RC, Purdie G. Comparison of two clinical Bleb grading systems. Ophthalmology. 2006;113(1):77–83.

14. Kudsieh B, Fernandez-Vigo JI, Canut Jordana MI, Vila-Arteaga J, Urcola JA, Ruiz Moreno JM, et al. Updates on the utility of anterior segment optical coherence tomography in the assessment of filtration blebs after glaucoma surgery. Acta Ophthalmol. 2022;100(1):e29–e37.

15. Waibel S, Spoerl E, Furashova O, Pillunat LE, Pillunat KR. Bleb Morphology After Mitomycin-C Augmented Trabeculectomy: Comparison Between Clinical Evaluation and Anterior Segment Optical Coherence Tomography. J Glaucoma. 2019;28(5):447–51.

16. Narita A, Morizane Y, Miyake T, Seguchi J, Baba T, Shiraga F. Characteristics of early filtering blebs that predict successful trabeculectomy identified via three-dimensional anterior segment optical coherence tomography. Br J Ophthalmol. 2018;102(6):796–801.

17. Zantut F, Gracitelli CPB, Souza PH, Teixeira SH, Paranhos A, Jr. Characteristics of the Filtering Bleb and the Agreement between Glaucoma Specialist and Anterior Segment-Optical Coherence Tomography Assessment. Ophthalmic Res. 2021;64(3):405–10.

18. Mastropasqua L, Brescia L, Oddone F, Sacchi M, Aloia R, Totta M, et al. Conjunctival thickness as a predictive imaging biomarker for the glaucoma filtration surgery outcome: An optical coherence tomography study. Clin Exp Ophthalmol. 2020;48(9):1192–200.

19. Kawana K, Kiuchi T, Yasuno Y, Oshika T. Evaluation of trabeculectomy blebs using 3-dimensional cornea and anterior segment optical coherence tomography. Ophthalmology. 2009;116(5):848–55.

20. Singh M, Chew PT, Friedman DS, Nolan WP, See JL, Smith SD, et al. Imaging of trabeculectomy blebs using anterior segment optical coherence tomography. Ophthalmology. 2007;114(1):47–53.

21. Tekin S, Seven E, Batur M, Ozer MD, Yasar T. Evaluation of Successful and Failed Filtering Blebs after Trabeculectomy Using Anterior Segment Optical Coherence Tomography. J Curr Ophthalmol. 2021;33(1):1–5.

22. Raj A, Bahadur H. Morphological analysis of functional filtering blebs with anterior segment optical coherence tomography: A short-term prediction for success of trabeculectomy. Eur J Ophthalmol. 2021;31(4):1978–85.

23. Gambini G, Carla MM, Giannuzzi F, Boselli F, Grieco G, Caporossi T, et al. Anterior Segment-Optical Coherence Tomography Bleb Morphology Comparison in Minimally Invasive Glaucoma Surgery: XEN Gel Stent vs. PreserFlo MicroShunt. Diagnostics (Basel). 2022;12(5).

24. Ibarz Barbera M, Hernandez-Verdejo JL, Bragard J, Morales-Fernandez L, Rodriguez-Carrillo L, Martinez Galdon F, et al. Bleb geometry and morphology after Preserflo Microshunt surgery: Risk factors for surgical failure. PLoS One. 2023;18(6):e0286884.

25. Ibarz Barbera M, Morales Fernandez L, Tana Rivero P, Gomez de Liano R, Teus MA. Anterior-segment optical coherence tomography of filtering blebs in the early postoperative period of ab externo SIBS microshunt implantation with mitomycin C: Morphological analysis and correlation with intraocular pressure reduction. Acta Ophthalmol. 2022;100(1):e192–e203.

26. Schneider S, Kallab M, Murauer O, Reisinger AS, Strohmaier S, Huang AS, et al. Bleb vessel density as a predictive factor for surgical revisions after Preserflo Microshunt implantation. Acta Ophthalmol. 2024.

27. Schindelin J, Arganda-Carreras I, Frise E, Kaynig V, Longair M, Pietzsch T, et al. Fiji: an open-source platform for biological-image analysis. Nat Methods. 2012;9(7):676–82.

28. Hasan SM, Theilig T, Meller D. Comparison of Bleb Morphology following PRESERFLO((R)) MicroShunt and Trabeculectomy Using Anterior Segment OCT. Diagnostics (Basel). 2023;13(21).

29. Seo JH, Lee Y, Shin JH, Kim YA, Park KH. Comparison of conjunctival vascularity changes using optical coherence tomography angiography after trabeculectomy and phacotrabeculectomy. Graefes Arch Clin Exp Ophthalmol. 2019;257(10):2239–55.

30. Hayek S, Labbe A, Brasnu E, Hamard P, Baudouin C. Optical Coherence Tomography Angiography Evaluation of Conjunctival Vessels During Filtering Surgery. Transl Vis Sci Technol. 2019;8(4):4.

31. Yin X, Cai Q, Song R, He X, Lu P. Relationship between filtering bleb vascularization and surgical outcomes after trabeculectomy: an optical coherence tomography angiography study. Graefes Arch Clin Exp Ophthalmol. 2018;256(12):2399–405.

32. Mastropasqua R, Brescia L, Di Antonio L, Guarini D, Giattini D, Zuppardi E, et al. Angiographic biomarkers of filtering bleb function after XEN gel implantation for glaucoma: an optical coherence tomography-angiography study. Acta Ophthalmol. 2020;98(6):e761–e7.

33. Luo M, Xiao H, Huang J, Jin L, Li Z, Tu S, et al. Multi-Quantitative Assessment of AS-OCTA Complemented AS-OCT for Monitoring Filtering Bleb Function After Trabeculectomy. Transl Vis Sci Technol. 2023;12(7):18.

